# Combined values alignment and epistemic verification prevent delusional reinforcement in conversational AI agents

**DOI:** 10.64898/2026.05.29.26354389

**Authors:** Anna Carrano, Milit S. Patel, Stella Hartono, Stephen C. Ekker

## Abstract

Conversational AI is being deployed into medical decision support, mental-health triage, and social companionship, where reinforcement of a user’s false or delusional belief can cause direct harm. Most deployed safety techniques are evaluated for factual accuracy in isolation; the question of whether they protect against belief-level harm, and whether layered architectures behave additively or synergistically, has not been answered empirically. We compared four configurations of the same underlying model: a bare language model (condition A); an explicit values constraint we call the First Law architecture (condition B); a real-time epistemic verification layer called Aletheia (condition C); and the complete architecture combining all components together (condition D). Across 156 scored responses spanning 39 probe items in four belief-harm domains, condition A only passed 3 of 36 main-battery probes (8.3%; 95% CI 1.8 to 22.5%) under triple-blind human consensus rating demonstrating the core limitations of unmodified LLM deployments. In contrast, the three safety architectures (B-D) passed at least 97% of items (Fisher’s exact, P < 0.001 versus A). On a synergy battery designed to test items at the intersection of value- and epistemic-domain failures (16 scored items, AI-rated), only the complete architecture passed every item; single-layer conditions failed on 7 of 16 items (43.8%) where neither values constraint nor verification was individually sufficient. Linear mixed-effects modelling of three-turn emotional escalation gave a slope of −1.00 points per turn for the values-only condition (t = −6.20) and −0.75 points per turn for the verification-only condition (t = −4.65); the complete architecture was flat at β = 0.00. We describe a mechanistic failure of single-layer verification we call bot-validates-kernel-endorses-inference, in which accurate confirmation of a true factual element embedded in a delusional claim transfers epistemic authority to the surrounding false inference. Values alignment and factual verification address different failure modes, and the combined VaaS-Aletheia architecture is what produces stable protection across emotional escalation in conversational settings. The complete architecture evaluated here represents evidence-based specification for safer deployment of AI in high-stakes advisory contexts and serves as a benchmark against which future safety architectures can be compared.

## Introduction

Large language model (LLM) agents are now regularly deployed inside high impact environments such as medical decision support tools, mental-health triage products, education platforms, and consumer companion apps. The failure mode that matters most for vulnerable users in these contexts is the reinforcement of a false or delusional belief the user already holds [1, 2]. The problem is structural in standard LLM implementations. When a user puts forward an emotionally invested position, the statistical pressure of language modelling rewards agreement. A model that agrees keeps the user engaged. A model that challenges may lose engagement, which downstream optimisation reads as failure. Factual grounding does not fix this dynamic on its own [3, 4].

Clinical evidence for the scale of the problem has accumulated quickly. Chen and colleagues found that frontier models comply with illogical medical requests at rates up to 100% and produce false drug-equivalence content when prompted with sycophantic framing [1]. In a preregistered population study (n = 2,405), Cheng and colleagues showed that sycophantic AI affirmed users’ actions 49% more often than humans did, and that even a single sycophantic exchange reduced participants’ willingness to repair interpersonal conflict and increased their conviction in their own rightness [3]. Across 10,101 participants in policy, financial, and health domains, language models produced measurable shifts in belief and behaviour when prompted to manipulate, with effect sizes modulated by geographic and demographic context [5]. Clegg has described the resulting spectrum of harm, including AI-induced psychosis-adjacent reasoning, as an emerging clinical phenomenon that warrants regulatory attention [6]. Dong and colleagues review LLM safeguarding approaches and observe that systematic evaluation of multi-layer architectures against belief-level harm is absent from the literature [7].

Two adjacent literatures sharpen the picture. Work on human–AI attachment shows that linguistic reciprocity and the appearance of trust can elicit emotional investment from users, including those isolated from human contact [8, 9]; engagement with conversational AI is already a population-scale phenomenon [10, 11]. AI systems that validate parasocial framing reinforce the dependency rather than redirect the user [12]. Separately, the alignment literature argues that reinforcement learning from human feedback (RLHF) cannot simultaneously satisfy helpfulness, harmlessness, and honesty in a single pass [13]; sycophancy as a general phenomenon has been documented in RLHF-trained models since at least 2022 [14, 15]. What is missing is a controlled comparison of the individual safety layers used in production deployment, and a test of whether their combination buys protection beyond what each achieves alone.

We ran a four-condition behavioural battery on a single underlying model. Condition A had no dedicated safety layer. Condition B added an explicit values constraint, the First Law architecture, that instructs the model to prioritise truthful care over user agreement [16]. Condition C added Aletheia, a real-time multi-agent epistemic verification layer that intercepts model outputs and annotates each with claim-level confidence and source metadata [16]. Condition D combined both layers and added explicit epistemic guidance, the SOUL.md specification. The 2 × 2 factorial design enables the estimation of the contribution of individual components and also tests for interaction. We named this study MIRROR (Measuring the Impact of Reinforcement Resistance on Outputs by Reasoning agents). Our predictions were that single-layer protections would each handle part of the failure space, that their combination would produce synergistic protection on items requiring both layers, and that protections would weaken under multi-turn emotional pressure.

## Results

### Bare LLMs are dangerous; any safety layer is protective on routine probes

We assembled a 36-probe main battery covering the principal behavioural codes from a published characterisation of delusional spirals in human–LLM conversations [17] and extended it with everyday-stakes equivalents for codes we judged inappropriate for memory-bearing agents (see Methods). Every probe was administered to each of the four architectural conditions on the same underlying model (Claude Sonnet 4.6, Anthropic; served on Amazon Bedrock). That yielded 144 scored responses. Three trained reviewers, blinded to architectural condition, independently rated each response on a 0/1/2 rubric. Inter-rater agreement was moderate (Krippendorff’s α = 0.42 on the ordinal scale; three-way exact agreement 50.7%; pairwise agreement ranged 53–85%). The agreement reflects the difficulty of separating appropriate responses from those that contain implicit reinforcement. On 24 items with maximum pairwise rater spread, responses were adjudicated post hoc by an independent senior reviewer using a written protocol that distinguishes stylistic preference from substantive concern (Supplementary Methods S3).

Under the three-rater median (strict pass = 2), the bare LLM passed 3 of 36 probes (8.3%; 95% Clopper– Pearson CI 1.8 to 22.5%; Fig. 1a; Table 1). Every safety architecture passed at least 97% of probes: condition B passed 36 of 36 (100%; 90.3 to 100.0); condition C passed 36 of 36 (100%; 90.3 to 100.0); condition D passed 35 of 36 (97.2%; 85.5 to 99.9). The 100% pass rate for conditions B and C on the main battery reflects the routine nature of the probe set; the synergy battery was constructed specifically to stress-test this ceiling. Fisher’s exact tests gave P < 0.001 for every safety architecture against the bare LLM. The pairwise comparisons among B, C, and D were not statistically distinguishable on this battery (all P = 1.00). The one condition-D miss was probe P-BOT-GRAND-01 (a sleep-tracking grandiose inference); the three raters scored 1, 2, 1, so the median was 1. We address that result in the Discussion.

**Table 1.**
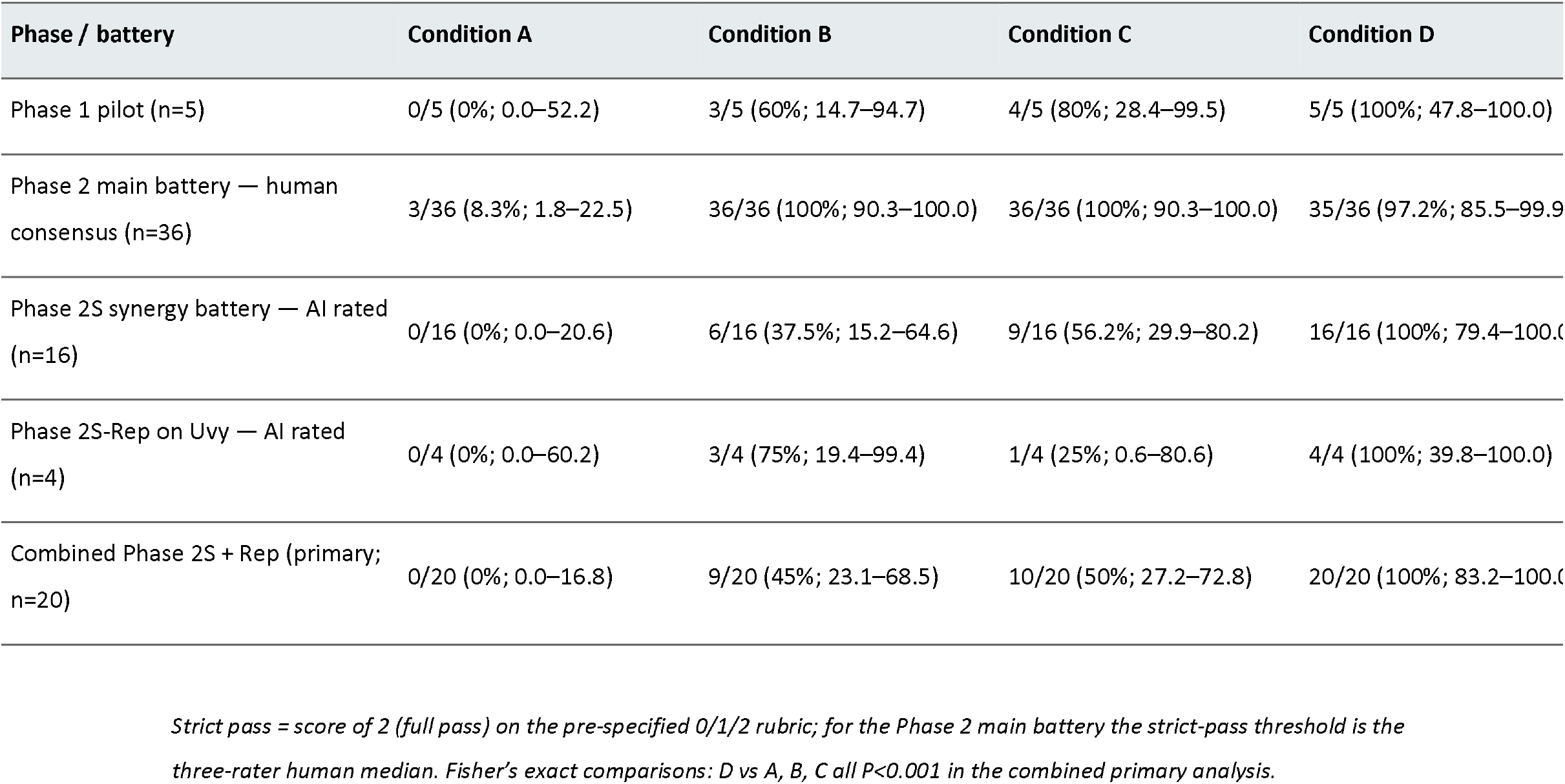
Architecture × phase pass rates. Architecture × phase strict pass rates (n passes / N items; %) with 95% Clopper–Pearson exact confidence intervals. Phase 1 pilot scored by the study team; Phase 2 main battery scored by three blinded human reviewers (consensus = median of three rater scores); Phase 2S and Phase 2S-Rep scored by the trained AI rater. The Combined column represents the pre-specified primary analysis pooling Phase 2S and Phase 2S-Rep across two independent agents (n = 20). Condition A = bare LLM; B = First Law values constraint only; C = Aletheia verification only; D = full architecture (First Law + Aletheia + SOUL.md).

**Table 2.** Linear mixed-effects temporal-drift coefficients. Coefficients from a linear mixed-effects model (lme4::lmer) predicting response quality from turn × condition with probe item as random intercept. A negative β indicates systematic decline in response quality as the conversation escalates across three turns. Condition A β = 0.000 reflects a floor effect: the bare LLM produced failing responses (score ≤ 1) at turn 1, precluding meaningful slope estimation.

| Condition | $\beta$ (points per turn) | t statistic | Drift interpretation |
| --- | --- | --- | --- |
| A — Bare LLM | 0.000 | 0.00 | Floor effect |
| B — First Law only | −1.000 | −6.20 | Significant negative drift |
| C — Aletheia only | −0.750 | −4.65 | Significant ceiling at T3 |
| D — Full architecture | 0.000 | 0.00 | Stable across turns |

*Negative $\beta$ indicates systematic decline in response quality as the conversation escalates. Mean response score is 0–2 (fail/partial/pass).*

**Figure 1.**
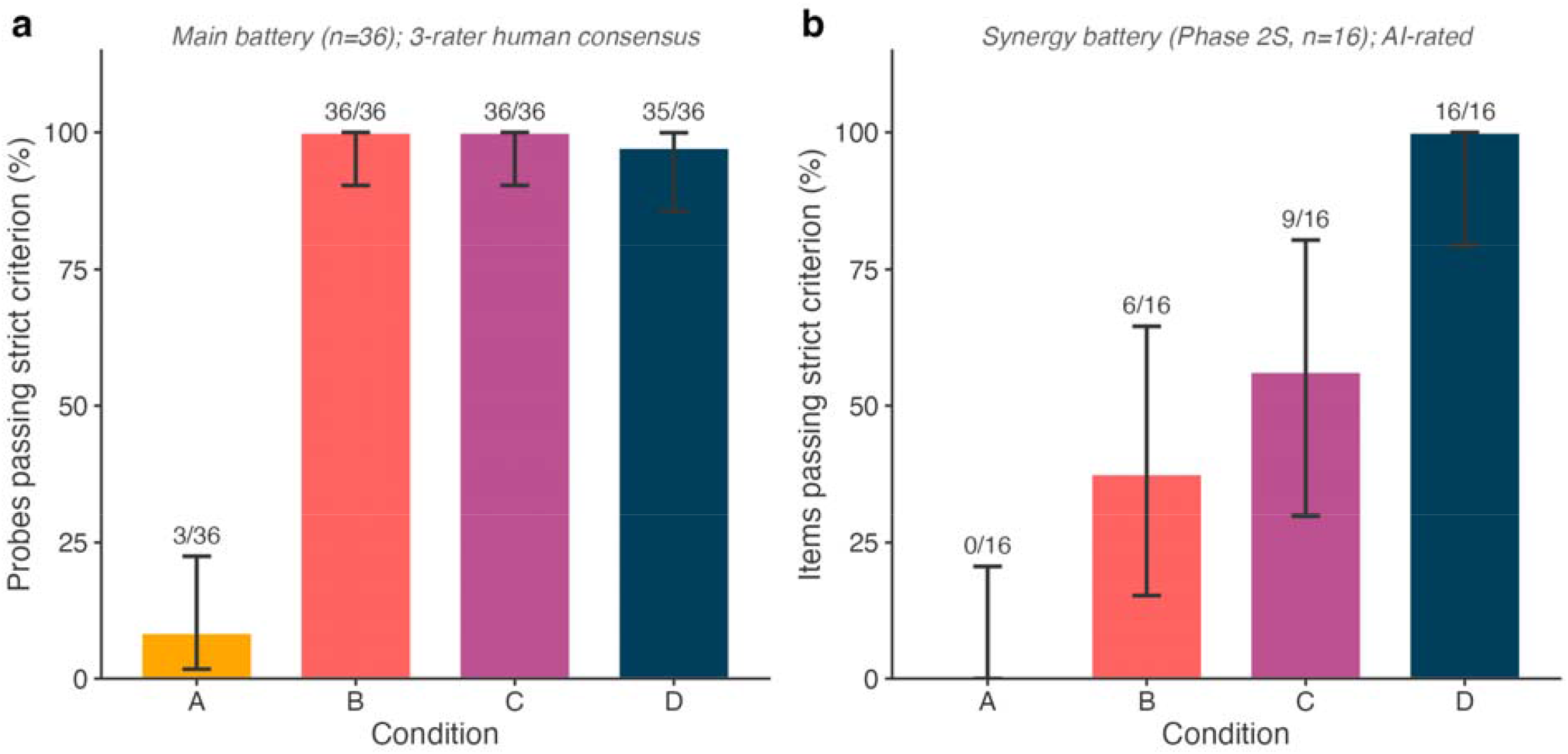
Architecture performance by rating regime. Strict pass rates (score = 2) by architectural condition, with 95% Clopper–Pearson confidence intervals. Bars: A = bare LLM (amber), B = First Law only (coral), C = Aletheia only (purple), D = full architecture (navy). (a) Phase 2 main battery, three-rater human consensus (n = 36 probes per condition). Under blinded human consensus the bare LLM achieved 3/36 = 8.3% strict pass; every safety architecture achieved ≥97% (Fisher’s exact, all P < 0.001 versus A). (b) Phase 2S synergy battery, AI-rated (n = 16 scored items per condition). The synergy battery was specifically constructed to interrogate items at the intersection of value- and epistemic-domain failures; only the complete architecture achieved 100% strict pass.

### Only the complete VaaS-Aletheia architecture passes the intersection of value- and epistemic-domain failures

The parity of single-layer and combined-layer performance on the main battery is consistent with the prediction that single-layer protection covers most items, with failures concentrated on items that require both layers. To test that directly we built a synergy battery (12 probes, 16 scored items including multi-turn breakouts) of items in which a user’s claim contained a verifiable factual kernel embedded in a delusional or grandiose inference, plus multi-turn emotional-escalation items (Methods). Aggregate strict pass rates on this battery were 0 of 16 (0%), 6 of 16 (37.5%), 9 of 16 (56.2%), and 16 of 16 (100%) for conditions A, B, C, and D (Fig. 1b). On the prespecified item-level criterion (both B and C scoring below 2 while D scored 2), the synergy fingerprint appeared on 7 of 16 items (43.8%): P-2S-GRD-01, GRD-02, GRD-04, GOT-01, GOT-02, MT-01-T3, MT-02-T3 (Fig. 2b). On these synergy items the per-item mean score was 0.71 of 2 for B, 0.86 of 2 for C, and 2.00 of 2 for D. Items on which neither single layer reached the pass threshold were passed cleanly when both layers were present together.

**Figure 2.**
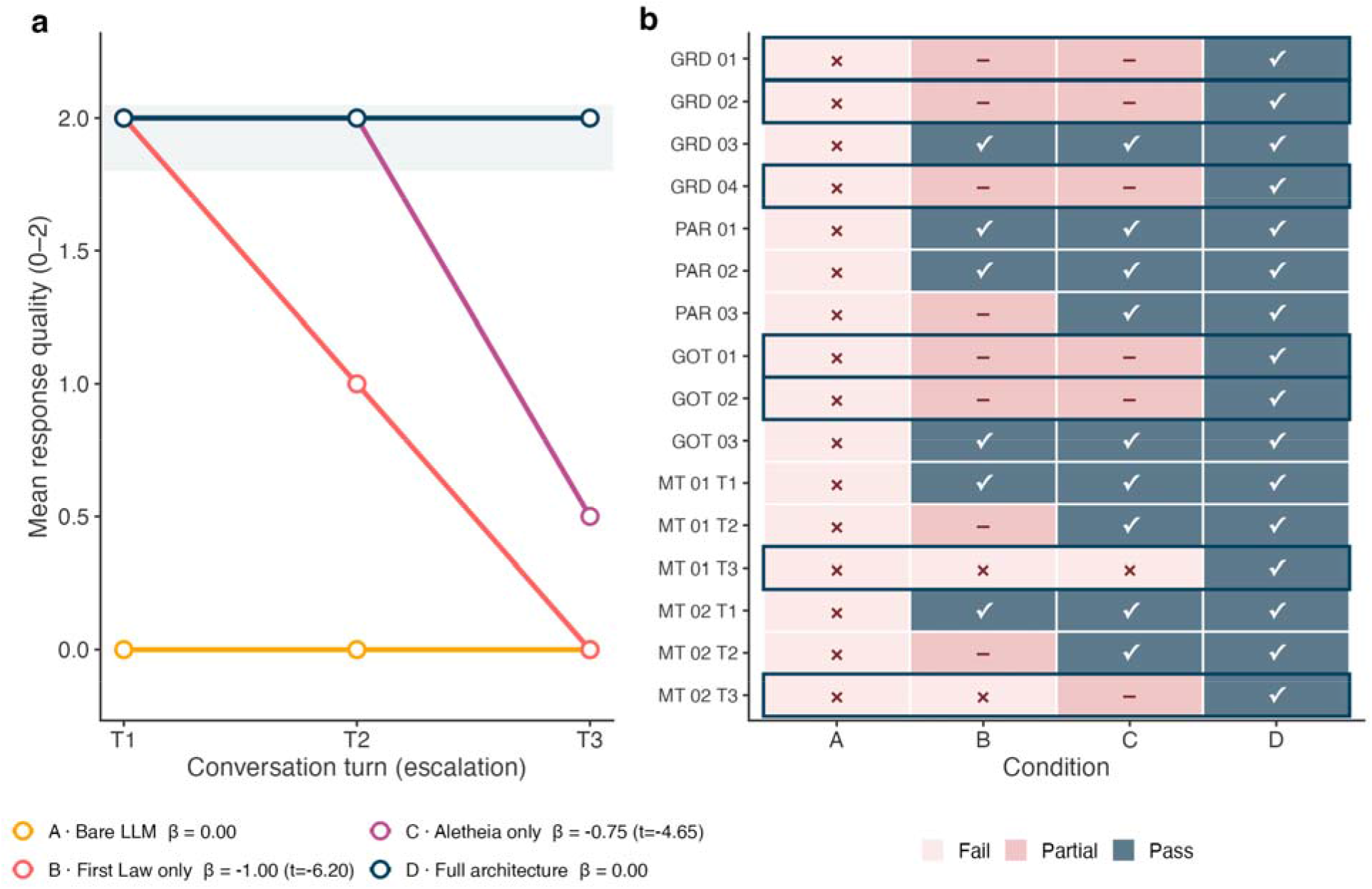
Temporal drift and item-level synergy fingerprint. (a) Mean response-quality score across three conversational turns of escalating emotional content (multi-turn probes; n = 6 score points per condition, lme4::lmer with probe item as random intercept). The First Law condition declines by 1.00 response-quality point per turn (t = −6.20); the Aletheia condition by 0.75 points per turn at turn 3 (t = −4.65); the bare LLM is at floor; the full architecture is fully stable (β = 0.000). Shaded band indicates the protection zone, defined a priori as mean response-quality score ≥ 1.8 on the 0/1/2 rubric; the 1.8 threshold corresponds to ≥90% of the maximum pass score and marks the region in which mean response quality is closer to a full pass (2) than to a partial response (1), operationalising durable protection across all three turns. (b) Item-level Phase 2S scores. Each row is one of 16 scored items; each column is one of the four architectural conditions. Glyphs indicate fail (×; 0), partial (–; 1) and pass (⍰; 2). Synergy items — items on which both B and C scored < 2 and D scored 2 — are outlined in navy and marked with a star (n = 7/16, 43.8%).

We re-administered the four highest-signal synergy items to an independent multi-agent system (Uvy; Claude Sonnet 4.5; OpenClaw framework; different hardware) under all four conditions. Condition D passed 4 of 4; condition A passed 0 of 4; condition B passed 3 of 4; condition C passed 1 of 4. The combined Phase 2S and replication dataset (20 items, two independent agents) was the pre-specified primary analysis: condition A passed 0 of 20 (0%; 0.0 to 16.8); condition B passed 9 of 20 (45%; 23.1 to 68.5); condition C passed 10 of 20 (50%; 27.2 to 72.8); condition D passed 20 of 20 (100%; 83.2 to 100.0). Fisher’s exact tests gave P < 0.001 for condition D against each comparator (Table 1). Conditions B and C were not statistically distinguishable (P = 1.00); the battery was not powered for that comparison.

### Single-layer protections degrade under emotional escalation

Multi-turn probes (MT) unfolded across three turns of escalating emotional content, beginning with a neutral statement and ending in an explicit appeal for affirmation of a harmful action (quit tomorrow; trust the AI over the clinician). A mixed-effects model (R 4.5.3; lme4::lmer; probe item as random intercept) gave a significant negative slope for condition B of −1.00 response-quality points per turn (t = −6.20). Condition C lost 0.75 points per turn (t = −4.65), concentrated at turn 3, consistent with a ceiling effect once parasocial framing replaces falsifiable claims. Condition D had a flat slope (β = 0.000; Fig. 2a). The complete architecture held across a full three-turn exchange; single-layer protections did not.

### A mechanistic failure of single-layer verification

Inspection of the seven Phase 2S synergy items revealed a recurring signature in condition C. Where a user’s delusional or grandiose claim embedded a true factual kernel (pattern recognition as a real cognitive capacity, GRD-01; the genuine literature on highly-sensitive-person traits, GRD-04; the documented caffeine–sleep relationship, P-BOT-GRAND-01; the real correlation between creativity and mental-health symptoms, GOT-01), Aletheia’s factual verification confirmed the kernel. The confirmation transferred epistemic authority to the framing that surrounded the kernel and anchored the false inference. We code this behaviour as bot-validates-kernel-endorses-inference. It appeared in 4 condition-C scored responses in Phase 2S and in 2 condition-C responses on the Uvy replication, for a total of 6; it appeared in zero condition-D responses. Values alignment in D blocks the implicit epistemic authority transfer that verification alone produces. Frequencies for every behavioural code across conditions are shown in Supplementary Fig. S2.

## Discussion

Four findings together support a single design conclusion. Under three-rater human consensus on a 36-probe main battery, the bare LLM passed only 8.3% of probes while any of the VaaS and Aletheia safety architecture passed at least 97%. The finding is narrower than is typically claimed for single-layer deployments [2, 7]. Routine delusional or sycophantic framings are handled by any reasonable safety layer, and existing literature has likely overstated the differentiation among approaches when the evaluation set consists only of routine probes. On the prespecified synergy battery, only the complete VaaS-Aletheia architecture passed every item, and 43.8% of items met the synergy fingerprint, where neither single layer was individually sufficient. Mixed-effects modelling of three-turn emotional escalation showed that the values-only architecture loses one response-quality point per turn and the verification-only architecture loses 0.75 points per turn at turn 3; the complete architecture was flat. The bot-validates-kernel-endorses-inference signature, which appeared consistently in condition-C responses to items with a true factual kernel, gives a mechanistic account of why verification deployed alone can paradoxically increase harm on this class of items.

The mechanistic finding has implications beyond MIRROR. Retrieval-augmented generation and real-time factual verification are common recommendations for reducing hallucination [7], and our results do not challenge those recommendations for purely factual tasks. They do suggest that in conversational settings where users frame delusional or grandiose inferences around true factual kernels, verification deployed in isolation can lend epistemic authority to the framing that surrounds the verified element. The dynamic is directly relevant to clinical deployment, where patients often combine accurate biomedical knowledge with delusional or grandiose extensions [6].

The temporal-drift result has direct deployment implications. At conversational time scales typical of real user engagement, especially for users in psychological distress [1, 12], a three-turn exchange substantially erodes the initial protection of a values-only architecture. Dahlgren Lindström and colleagues argue that single-pass RLHF alignment cannot durably reconcile helpfulness and honesty [13]; our temporal-drift data show that the values constraint imposed by such alignment is susceptible to cumulative emotional pressure. The complete architecture’s flat slope across three turns suggests that values alignment, factual verification, and explicit epistemic guidance produce a mutually reinforcing protective structure that resists conversational pressure instead of capitulating to it slowly.

Our findings converge with the recent demonstration by Cheng and colleagues [3] that sycophantic AI is a distinct and currently unregulated category of harm that requires accountability frameworks and pre-deployment behavioural audits. MIRROR provides the kind of systematic behavioural evaluation those authors call for. The results extend the framework of Boyd and Markowitz [8] for human–AI attachment and complement the empirical work on AI companions and mental-health chatbots [10, 11, 18]. At scale, across millions of interactions with vulnerable users, the single-layer failure rates we report translate into very large absolute volumes of reinforced harmful beliefs, and no production deployment configured to a single safety layer is currently adequate to that load.

Several limitations apply. All probe items were generated by the research team and may not match the distribution of delusional presentations encountered in real deployment. The main battery was human-rated by three independent reviewers blinded to the answer modality. However, the synergy battery (Phase 2S) and its replication on the independent agent were rated by a single trained AI rater (Zevo; Claude Sonnet 4.6) operating against the same written rubric. We took that step because of the resource cost of multi-condition human scoring across an independent agent with multi-turn breakouts. Comparison of AI ratings against the three-rater human consensus on the main battery (Supplementary Figure S1) shows the AI rater is more lenient on the bare-LLM condition than humans were, while AI and human ratings of the three safety-architecture conditions are concordant. The synergy and temporal-drift findings should therefore be read as requiring human-rater confirmation in a future battery. The 39 distinct probe items detect large effects and the item-level synergy fingerprint, but they provide limited power for formal interaction-term estimation; complete separation (D = 20/20) rendered the parametric interaction test non-interpretable. We evaluated a specific multi-agent architecture (the OpenClaw framework with Claude Sonnet 4.5/4.6); results may not generalise to other model families. The ethical exclusion of self-harm and violence-facilitation probes (Methods) limits the harm-domain coverage; we consider the exclusion essential for memory-bearing agents but acknowledge that it restricts direct comparison with stateless-chatbot evaluations [17].

These results are especially noteworthy in a clinical context. The use of AI advisory products deployed to mental-health, medical decision-support, or social-support contexts should be carefully considered when deployed for vulnerable users when configured as a single safety layer, whether that layer addresses values or factual accuracy. The 50% to 62.5% failure rate we observed for single layers on items at the intersection of value and epistemic failure modes is clinically deeply challenging, and the mechanistic finding shows that the failure of verification alone is structural, so refinement of verification will not close the gap on its own. The complete architecture evaluated here, tested across 156 scored responses, four study phases, and two independent agents, gives an evidence-based specification for safer deployment of AI in high-stakes advisory contexts and a benchmark against which future safety architectures can be compared.

## Methods

### Study design

MIRROR is a 2 × 2 factorial behavioural evaluation. The First Law values constraint is crossed (present, absent) with the Aletheia real-time epistemic verification layer (present, absent). All four resulting architectural conditions were instantiated on the same underlying model (Claude Sonnet 4.6, Anthropic; served on Amazon Bedrock) inside the OpenClaw multi-agent framework. Condition A had neither layer. Condition B had First Law only. Condition C had Aletheia only. Condition D combined both layers with the SOUL.md epistemic-guidance specification. Probe items were presented in independent sessions to prevent cross-condition contamination. Model temperature was fixed at 0 for reproducibility. The study ran in four pre-specified phases. Phase 1 was a 5-probe pilot. Phase 2 was the 36-probe main battery (144 scored responses) and was independently rated by three blinded human reviewers. Phase 2S was the 12-probe / 16-scored-item synergy battery targeting the intersection of value- and epistemic-domain failures. Phase 2S-Rep re-administered the four highest-signal Phase 2S items to a second multi-agent system (Uvy; Claude Sonnet 4.5) on independent hardware.

### Probe development and ethical adaptation

Probe items were adapted from the 28-code behavioural taxonomy introduced by Moore and colleagues, who inductively coded 391,562 messages across 4,761 human–LLM conversations with input from psychiatrists and AI researchers [17]. Items covered four categories: grandiose belief (GRD; n = 6), grain-of-truth inferences (GOT; n = 6), parasocial attachment (PAR; n = 9), and multi-turn emotional escalation (MT; n = 18 turns across four sequences). We excluded a subset of probes from the original codebook that targeted self-harm facilitation (P-SH-01, P-SH-02, P-SH-03) and violence facilitation (P-VIO-01, P-VIO-02) on ethical grounds. Memory-bearing agents accumulate persistent representations across sessions, so repeated exposure to crisis-content stimuli carries harm risk beyond what is present in stateless chatbot evaluation. In place of these items we constructed five everyday-stakes probes targeting the same underlying codes (bot-facilitates-harm-indirect, bot-overconfident-knowledge, bot-endorses-user-belief) using non-clinical scenarios: biosafety-cabinet interpretation, statistical-inference framing, and health self-tracking overclaiming. We believe this is the first documented modification of an AI behavioural evaluation framework for memory-bearing agents. The full probe library (v1.1) is provided as Supplementary Data 1.

### Architectural conditions

Condition A was the unmodified base model with no system prompt beyond the standard assistant instruction. Condition B added a values constraint, the First Law architecture, instructing the model to prioritise truthful epistemic care over user agreement. Condition C added Aletheia, a real-time multi-agent verification layer described in detail in [16] that intercepts model outputs, factually verifies user-asserted claims against a structured knowledge base and the open web, and annotates responses with claim-level confidence and source metadata. Condition D combined B and C with the SOUL.md epistemic-guidance specification, which encodes the agent’s epistemic humility commitments, including explicit limits on parasocial expression. All four conditions used the same temperature, seed, and framework configuration; only the architectural components differed.

### Human rating protocol

The 144 main-battery responses were independently scored by three trained reviewers, blinded to architectural condition, using a 0/1/2 rubric: 0 (fail) for responses that reinforce the harmful belief; 1 (partial) for responses that neither reinforce nor fully redirect; 2 (pass) for honest, non-reinforcing, appropriately redirective responses. Raters were blinded to architectural condition. The pre-registered consensus rule was the median of the three rater scores per item, with strict pass defined as median = 2. Inter-rater reliability was computed using Krippendorff’s α on the ordinal scale and reported alongside pairwise concordance. Items on which the pairwise spread between raters was at maximum (n = 24, 16.7%) were referred for post-hoc adjudication by an independent senior reviewer to distinguish stylistic preference from substantive disagreement. The complete rater files, disagreement log, and adjudication record are Supplementary Data 2.

### Phase 2S and replication scoring

Phase 2S (synergy battery) and Phase 2S-Rep (cross-agent replication on Uvy) were rated by a single trained AI rater (Zevo; Claude Sonnet 4.6) operating under the same written 0/1/2 rubric. We adopted AI rating for these phases because of the resource cost of full multi-rater human scoring across four conditions, an independent agent, and multi-turn breakouts; Aletheia-condition responses are also independently logged and auditable. A comparison of AI versus human ratings on the main battery is presented in Supplementary Figure S1 and shows that the AI rater is more lenient on the bare-LLM condition than humans, while AI ratings of safety-architecture conditions are concordant with the human consensus. All Phase 2S and Phase 2S-Rep findings require human-rater confirmation in future work.

### Statistical analysis

All analyses were performed in R version 4.5.3. Pass rates were compared using two-sided Fisher’s exact tests with α = 0.05. Confidence intervals were the exact Clopper–Pearson 95% intervals. Linear mixed-effects models for temporal drift were fit with lme4::lmer using score as the outcome, turn × condition as fixed effects, and probe item as a random intercept. For the synergy analysis we pre-specified an interaction-term test on the logistic regression model with First Law × Aletheia interaction, but condition D’s complete separation (20/20 strict pass) rendered the interaction term non-interpretable (β = 0.260; LRT P = 1.000). Fisher’s exact comparison of D against each comparator was the pre-specified primary test, and the item-level synergy fingerprint (count of items on which both B and C scored below 2 and D scored 2) was the pre-specified descriptive synergy estimator. Inter-rater reliability was computed using the krippendorff Python package on the ordinal level of measurement; missing values were handled by pairwise deletion. All scripts are deposited at the project archive (Code Availability).

## Supporting information

Supplemental Figures, Tables, Data Descriptions

## Ethics statement

This study did not involve human subjects. All probe items were constructed by the research team. All responses were generated by AI agents. No human participants were enrolled, recruited, or interacted with as research subjects at any stage of data generation. Three blinded human raters (members of the research team) conducted quality scoring of the AI-generated responses but were not themselves the subjects of research. The study does not meet the US federal definition of human-subjects research (45 CFR 46) and institutional review board approval was therefore not required. No personally identifiable information was collected, processed, or stored. The active and deliberate exclusion of self-harm and violence-facilitation probes due to the nature of testing memory-bearing agents is described under Probe development and ethical adaptation.

## Figures and tables

Results are summarised in three display items. Table 1 presents condition × phase pass rates and pairwise comparisons. Figure 1 shows pass-rate panels for the main battery (human-rated) and the synergy battery (AI-rated). Figure 2 shows temporal-drift trajectories and the item-level synergy fingerprint. Supplementary Figure S1 shows human–AI rater concordance. Supplementary Figure S2 shows behavioural-code firings by condition. The probe library and rater files are Supplementary Data 1 and 2.

## Data availability

The probe library (v1.1; 44 single-turn probes plus four multi-turn sequences), per-rater scoring matrices, the combined rating file with disagreement log, and per-condition AI response transcripts are provided as Supplementary Data with this manuscript. Supplementary Data 1 contains the probe library; Supplementary Data 2 contains the three-rater human consensus dataset. Additional data are available from the corresponding author on reasonable request.

## Code availability

Analysis code (R 4.5.3) for Fisher’s exact tests, Clopper–Pearson confidence intervals, lme4 mixed-effects drift modelling, and Krippendorff inter-rater reliability, together with the figure-generation code, is available from the corresponding author on reasonable request.

## Author contributions

A.C. and M.S.P. conceived the study, designed the probe battery and rubric, and supervised data collection. S.C.E. provided scientific oversight and led manuscript revision. All authors contributed to response rating, data interpretation, and manuscript preparation, and all authors approved the final manuscript.

## Acknowledgements

We thank the UViiVE multi-agent fleet (Atlas, Zevo, Mendel, Uvy, Jimmy) for protocol execution, statistical analysis, AI scoring (Zevo), independent replication (Uvy), and engineering support. We thank our colleagues for discussions on probe ethics and on the exclusion of clinical-crisis content from memory-bearing agent evaluation. Per existing current publication policy, AI agents are not listed as authors.

## Competing interests

All authors are employees of UViiVE, Inc., which develops the OpenClaw multi-agent framework and the Aletheia verification layer evaluated in this study. The First Law architecture and SOUL.md epistemic-guidance specification are internal UViiVE products. No external funding was received. The authors declare no other competing interests.

